# The remarkable ups and downs of birth rate in Switzerland 2020 to 2023 in a historical context

**DOI:** 10.1101/2023.12.05.23299432

**Authors:** Mathilde Le Vu, Katarina L. Matthes, Kaspar Staub

## Abstract

We follow population trends in the monthly birth rate in Switzerland almost up to the present and place the latest developments in a historical context. Birth rates in 2022 were the lowest since the 1870s, and the trend is continuing in 2023. The latest decline had already begun 1-2 years before Covid-19. Previous pandemics (1890, 1918, 1920, 1957) had led to a temporary decline in births ∽9 months after the epidemic peaks. With Covid-19, this appears more complex. During and shortly after the first two waves and shutdowns in 2020, there were more conceptions and thus excess birth rates in 2021, in all available subgroups except Italian-speaking Switzerland, and somewhat more pronounced among >30-year-old mothers and second parities. Possible reasons for the mini-boom include: The increased time at home during the shutdowns has – planned or not – led to more conceptions which has brought pregnancies forward; the corona virus was still circulating too infrequently in this 1st phase of the pandemic to have a negative impact on pregnancies or fertility; at the end of the waves and shutdowns, the perceived end of the pandemic threat could have led to an optimistic mood and thus also more conceptions. The subsequent decline from January 2022 was stronger than the increase before. The first part of the decline in 2022 is most likely due to a negative rebound from the advancement of births in 2020/2021 and deliberately postponed pregnancies due to the start of the vaccination program. The second part of the decline in 2022 is associated with conceptions during the large Omicron wave in the winter of 2021/2022, when many people in Switzerland fell ill. In addition, prices have been rising and real wages falling since 2021, the global political situation has become more unstable, and a general change in values regarding the willingness to have children may also be underway. Following these observations at population level (with limited depth of variables), more in-depth studies must now follow to better understand the dynamic ups and downs in the birth rate in Switzerland in recent years.

## Introduction

At the end of 2023, birth rates are the focus of attention in many places (Danny, 2023). Were there more or fewer births during or after the COVID-19 pandemic crisis, a baby boom or a baby bust? A multinational study showed that in many European countries, the initial pandemic shock in early 2020 was associated with a decline in births nine months later (Sobotka et al., 2023). Subsequently, some European countries reported stable or slightly increasing birth rates in 2021, nine months after the first and second pandemic waves in 2020. From January 2022, many European countries show a marked decline in birth rates, continuing a negative trend that may have begun before Covid-19 (Sobotka et al., 2023). In the US, the pattern of birth rates was somewhat different, with an initial decline in 2020 to early 2021 (Bailey et al., 2023), and especially after the first and second waves of Covid-19 (Adelman et al., 2023), followed by a smaller than expected rebound in 2022.

Birth rates are known to respond to pandemics and other crises (Lee et al., 2023), including heatwaves and natural disasters such as tsunamis (Barreca et al., 2018; Nobles et al., 2015). In the case of pandemics and epidemics, it has been shown that birth rates appear to drop approximately 9 months after the peak of an outbreak (as was the case with SARS-CoV-1, Zika, and to a lesser extent Ebola), followed by a rebound in births (Pomar et al., 2020). The reasons for this pattern are multifactorial but are likely to be related to deliberate postponement of conception, as well as illness-related fertility restrictions and natural abortions early in pregnancy during the peak of the outbreak. Most of the evidence on historical pandemics comes from research on the 1918-1920 influenza pandemic (“Spanish flu”), when births declined 9 months after the pandemic peak in Scandinavia (Bloom-Feshbach et al., 2011; Svenn-Erik Mamelund, 2004; Pomar et al., 2020), Britain (Chandra et al., 2018; Chandra & Yu, 2015; Reid, 2005), Japan (Chandra & Yu, 2015), and the United States (Bloom-Feshbach et al., 2011; Chandra et al., 2018). However, some of these aspects are currently being debated in the literature, for example whether the 1918-1920 pandemic or the end of the First World War is more likely to be associated with these changes in birthrates (Gaddy & Ingholt, 2023; S.-E. Mamelund, 2012).

Following the sharp decline in the birth rate in Switzerland from the 1970s onwards with the introduction of the contraceptive pill, the birth rate has stabilized at a low level in recent decades. The birth rate slightly declined for two or three years before the outbreak of the COVID-19 pandemic. Switzerland was hit by the first wave of COVID-19 in the spring of 2020, leading to a partial shutdown from mid-March to early May 2020. The stronger second wave emerged from October 2020 and led to historic excess mortality (Staub et al., 2022), especially among the elderly, accompanied again by a partial shutdown from November 2020 to February 2021, and followed by a gradual easing of these measures. As elsewhere, the circulation of different SARS-CoV-2 strains led to several waves of various intensity from 2021. The large omicron wave, which occurred between November 2021 and April 2022, resulted in very high case numbers but a relatively low mortality. Switzerland was also directly or indirectly affected by other events that coincided with the pandemic, such as the outbreak of war in Ukraine in February 2022 and a severe heat wave in the summer of 2022. In addition, COVID-19 led to an economic downturn.

For Switzerland, the most recent trends in birth rates in the years 2020 to 2023 in the context of the COVID-19 pandemic and crisis have not yet been scientifically studied. The aim of the present study is to describe the monthly dynamics in birth rates during the last two to three years of the COVID-19 pandemic, also for certain subgroups, and to place these population-level developments in a larger temporal context since the end of the 19th century.

## Data and methods

For this study, we have chosen two different approaches to the official Swiss vital statistics BEVNAT of the Swiss Federal Statistical Office (FSO). BEVNAT is the annual statistics of all births, marriages, divorces, deaths, etc. in Switzerland. On the one hand, we worked with the officially published monthly aggregated numbers of all live births in Switzerland between January 1871 and September 2023. The numbers for January through September 2023 are preliminary (final numbers for the full year 2023 will not be available until later in 2024). On the other hand, we obtained from the FSO all currently available individual data on all births per month from BEVNAT, in fully anonymized form and after signing a data contract (according to the Human Research Act HRA, no ethical approval is required when working with fully anonymized government data). Here we cover the period from January 1989 to December 2022 and have other co-factors such as maternal age, maternal nationality (Swiss vs. non-Swiss), parity at birth (since 2005), sex and vital status of the new-borns, and whether it was a multiple birth or not. We also know the language region (German-, French- or Italian-speaking Switzerland) in which the mother lived at the time of the birth. Language region can be considered a rough proxy for cultural, social and behavioral factors, including diet, smoking and alcohol consumption of the parents, as well as genetics (Novembre et al., 2008; Skrivankova et al., 2019). The timing and duration of the earlier pandemics of 1890, 1918-1920, 1957, 1969-1970 and 2009 were obtained from the weekly bulletin of the Federal Office of Public Health (Schweiz Bundesamt für Gesundheitswesen, n.d.).

Birth rates were calculated using the total annual population (source: FSO STATPOP). In a sensitivity analysis, we also calculated all models using the population of women aged 15-49 as the denominator. The monthly expected birth rate was estimated by using a Bayesian hierarchical model in which the last 5 years of the respective birth year are used for the calculation. A negative binomial model was used to model the number of births while the population was considered as an offset. Time and seasonality effects were added in the model as random effect using random walk model of order 1 and birth month as fixed effect. Following the model fitting process, we draw 1000 samples from the posterior distribution (the expected number of births). Subsequently, we computed the median and 95% credible intervals (CrI). The over-birth and under-birth rates were then determined by subtracting the expected values from the observed ones and expressing the results as percentages. These estimates are calculated for all subgroups mentioned above. In addition, using the same Bayesian hierarchical model, we predicted the birthrate from 2021-2023 using the years 2016-2020 (the pre-pandemic period) as reference birth numbers. All statistical analyses were performed using R Version 4.2.2 (R Core Team, 2022) and all models were fitted using INLA (Integrated Nested Laplace Approximation) (www.rinla.org).

## Results

The long-term trend in the monthly birth rate is shown in Figure 1. From the end of the 19th century at the latest, rates fell sharply to below 25 live births per 10 000 population. There was a temporary dip during the First World War. From the Second World War until the mid-1960s there was a marked increase in births due to the so-called baby boom generations. With the introduction of the contraceptive pill, the birth rate fell sharply from the 1970s and has remained below the fertility replacement level of 2.1 children per woman. In recent decades, the birth rate has more or less stabilized at a low level. After a slight increase in the birth rate around 2016, the birth rate declined slightly thereafter and until 2019 (Figure 2). In 2021, the second year of the pandemic, there was a temporary excess of births before the birth rate reached an all-time low of 9.3 live births per 10 000 population in 2022 (Supplementary Table S1). Preliminary monthly data up to September 2023 suggest that the downward trend is continuing (Figure 2).

**Figure 1:**
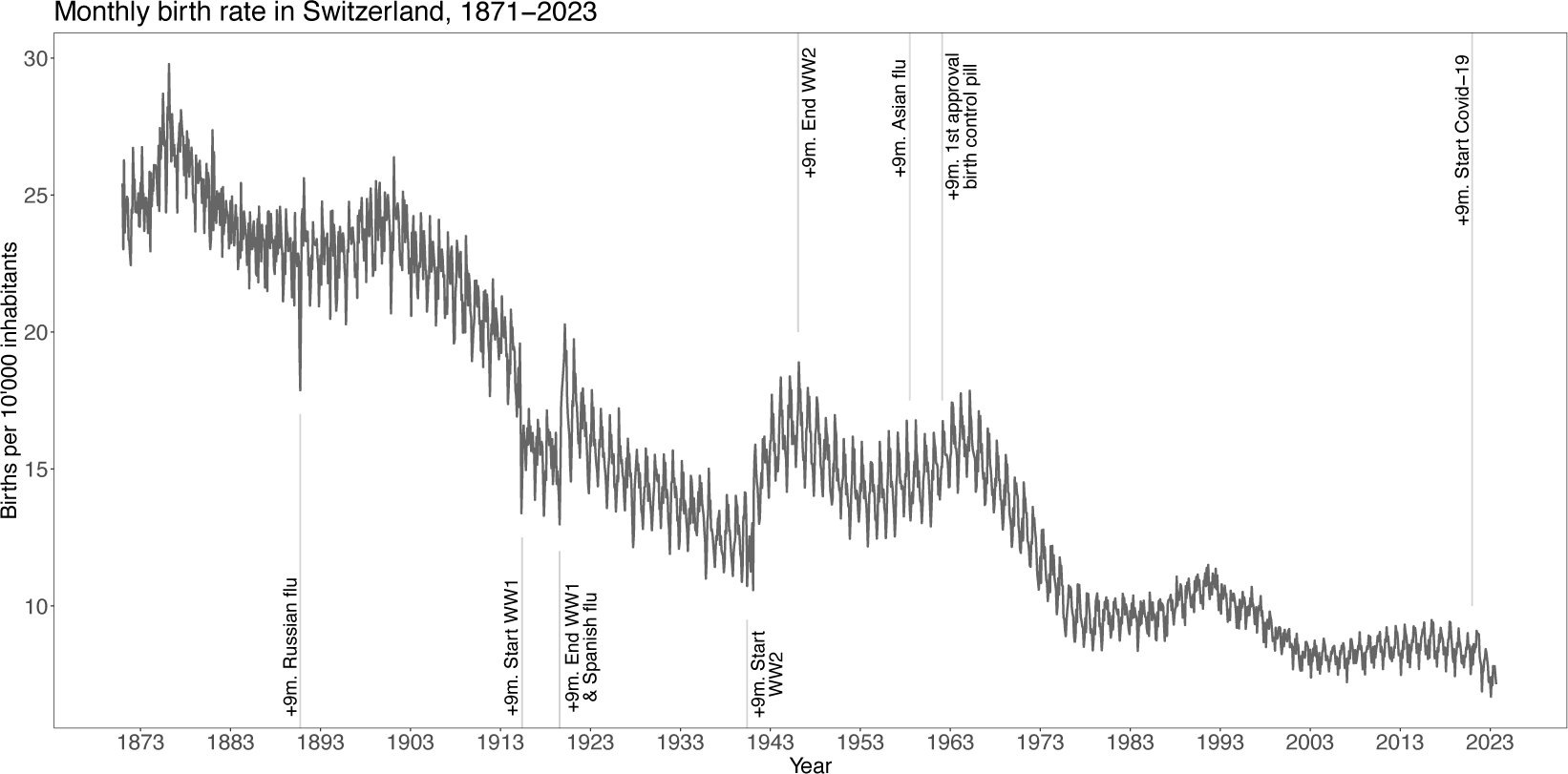
The long-term trend in the monthly live birth rate in Switzerland from 1871 to 2023. The annual data, also in relation to women aged 15-49 and the number of stillbirths, can be found in Supplementary Ta-ble S1. The monthly data for January to September 2023 are still provisional. (+9m. = 9 months after…)

**Figure 2:**
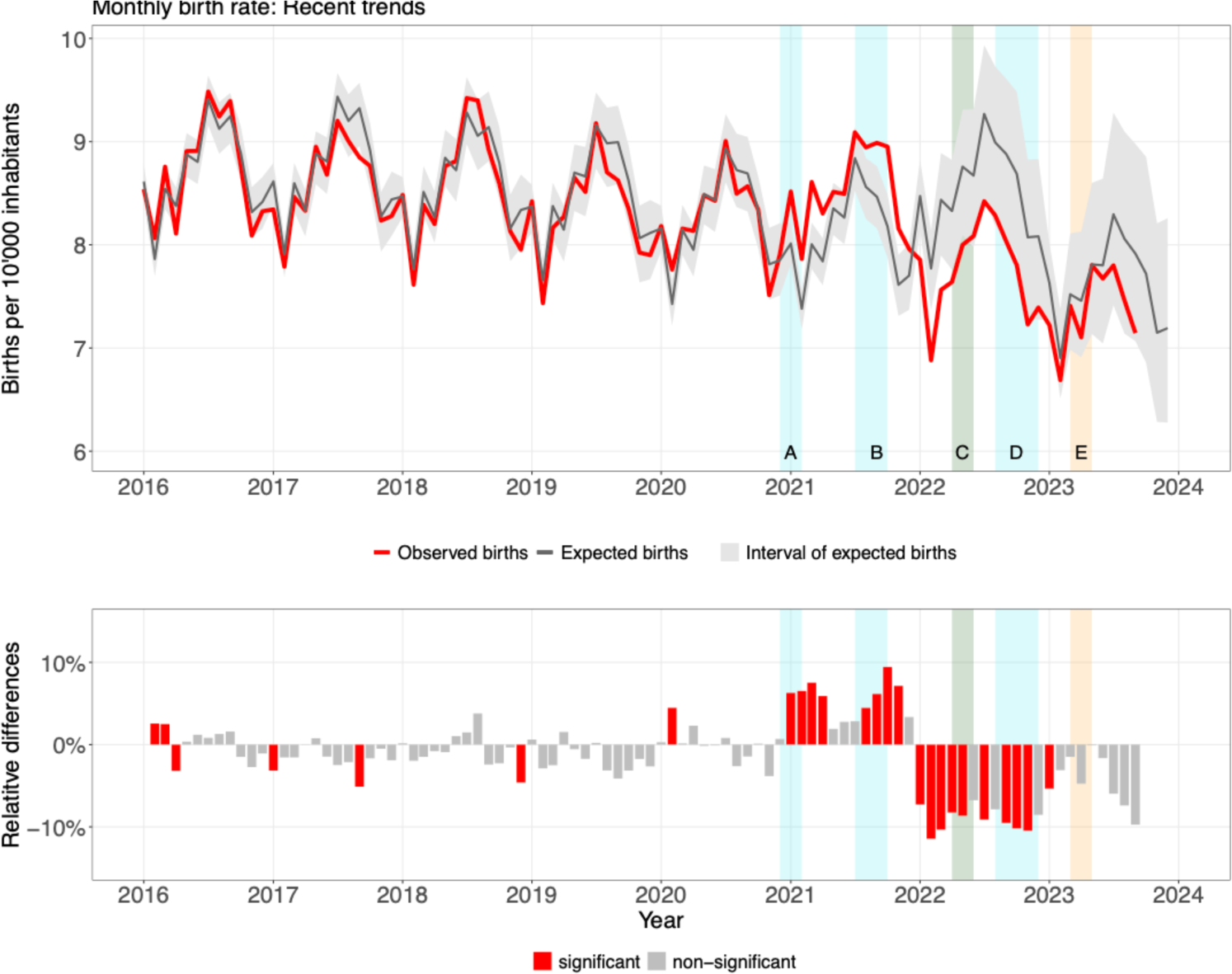
The monthly live birth rate in Switzerland from 2016 to 2023. Top: The observed values (red) compared with the expected values (grey, with interval). Below: The same numbers converted into relative differences, expressed in per cent (red=excess and lower birth rates). A: The time period 9 months after the first COVID-19 wave in Spring 2020 and the associated partial shutdown; B: The time period 9 months after the second COVID-19 wave in Fall and Winter 2020/2021 and the associated partial shutdown; C: The time period 9 months after the share of double-vaccinated young persons aged 20-40 years surpassed 30% to 50%; D: The time period 9 months after the large Omicron wave in Winter 2021/2022; E: The time period 9 months after the heat wave in Summer 2022.

If we look at the impact of previous pandemics on the birth rate, we find different patterns. The so-called “Russian flu”, which peaked in January 1890 and affected more than two thirds of the population, led to a decline in births of −21.8% (95% CrI −23.9 to −19.0%) about nine months later, in October 1890 (Supplementary Figure S1). About 9 months after the outbreak of World War I and the general mobilisation of troops in Switzerland in 1914, fertility rates began to fall and remained low for the duration of the war (Supplementary Figure S2). The first two waves of the “Spanish flu” occurred in 1918, at the end of the war, when birth rates had already fallen markedly. We see the lowest birth rates of the entire war period in mid-1919 (a decrease of −17.1% (95% CrI −34.9 to 6.2%)), about 9 months after the strongest wave of “Spanish flu”. About 9 months after the end of the war and the end of the second “Spanish flu” wave, there was a marked increase in births, which was temporarily interrupted again (by a decrease of −18.3%) towards the end of 1920, i.e. about 9 months after the strong later pandemic wave, which peaked in February 1920. The next influenza pandemic hit Switzerland in the autumn of 1957. Again, about 9 months after the peak of the “Asian flu”, we see a decrease in the birth rate of −12.1% (95% CrI −16.44 to −8.43%) (Supplementary Figure S3). The next influenza pandemic was the “Hong Kong flu”, which also hit Switzerland in the winters of 1968/69 and 1969/70, albeit to a lesser extent. This time we see no effect on the birth rate 9 months later (Supplementary Figure S4). The Great Recession of 2008/2009 and the “Swine Flu” pandemic at the end of 2009 also did not lead to a reduction in births 9 months later, but in the case of the “Swine Flu” an increase in births can be seen around 9 months after the end of the pandemic wave (Supplementary Figure S5).

If we look at the trend in recent years (Figure 2 and Table 1), there is no change at the population level in Switzerland around 9 months after the declaration of the COVID-19 pandemic and the first immediate pandemic shock at the beginning of 2020 compared to the expected values. On the other hand, we find a +4.5% to +9.4% increase in the monthly birth rate in two periods in 2021, associated with conceptions during and shortly after each of the two pandemic waves and the associated partial shutdowns in 2020. From January 2022, the relative differences in the birth rate compared to the expected monthly values are negative (−7.3% to −11.5%). The first months of this decline are associated with conceptions in the first quarter of 2021, when the vaccination campaign officially started in Switzerland. However, young adults without pre-existing conditions were not eligible for vaccination until May 2021, and among 20–40-year-olds, the proportion of double vaccinated people did not exceed 33% until July 2021 and 50% until September 2021 (this would correspond to births in April and June 2022). The second half of the decline in 2022 is associated with conceptions in the winter of 2021/2022, when the first major Omicron wave raged in Switzerland. The negative trend also continues in the provisional monthly data for 2023. A sensitivity analysis with the 15–49-year-old women as the denominator instead of the total population reaches very similar results. The monthly trend in the sex ratio at birth has been stable for many years and has not changed noticeably in recent years (Supplementary Figure S6).

**Table 1:**
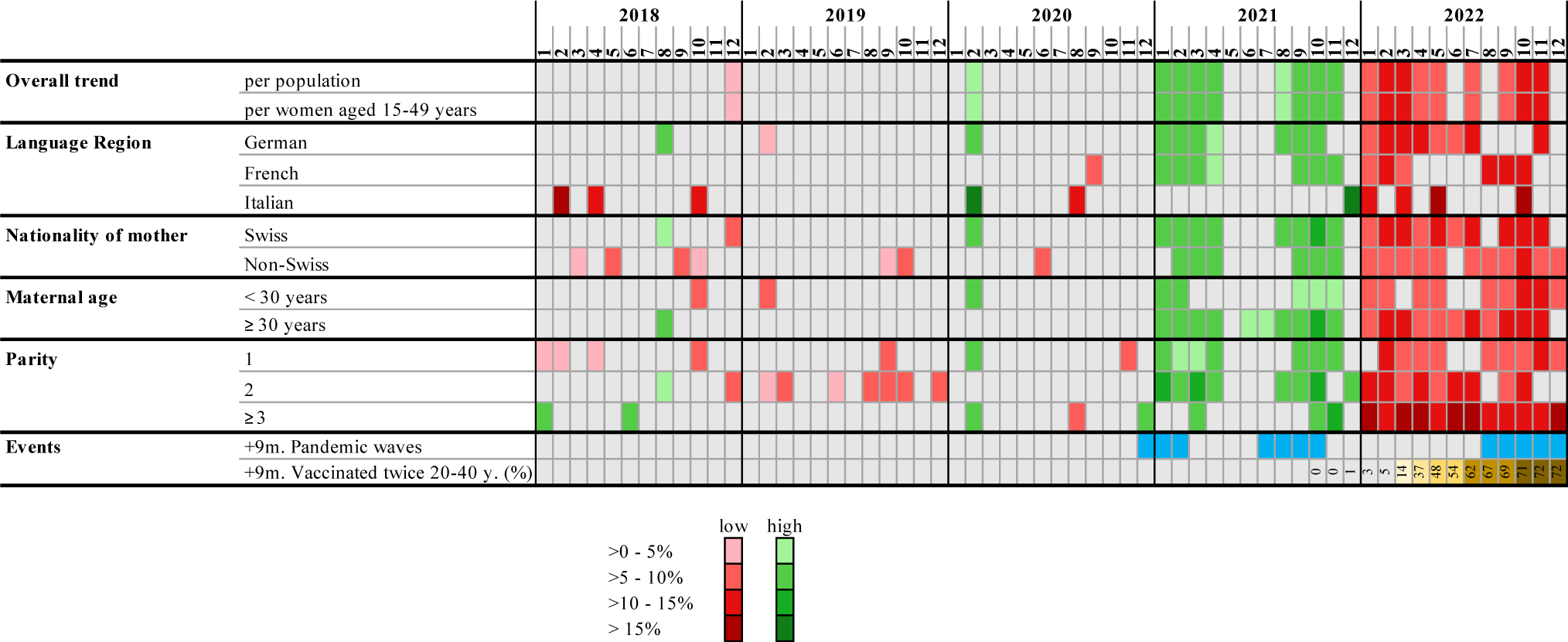
The monthly live birth rate in Switzerland from 2018 to 2022, shown as relative differences (expressed in percent) between observed vs. expected values for the overall trend and for available subgroups based on individual data. Excess and lower births s are color-coded in red or green.

If the increase in births in the year 2021 is not considered when estimating expected birth rates for 2022 and 2023, it becomes clear that the declining births in 2022 and 2023 follow the general trend of declining births that already began in the years before the pandemic (Figure 3). The observed births 2022 to 2023 are slightly lower than the expected births, but still follow the negative trend of birth rates predicted from the years 2016 to 2020.

**Figure 3:**
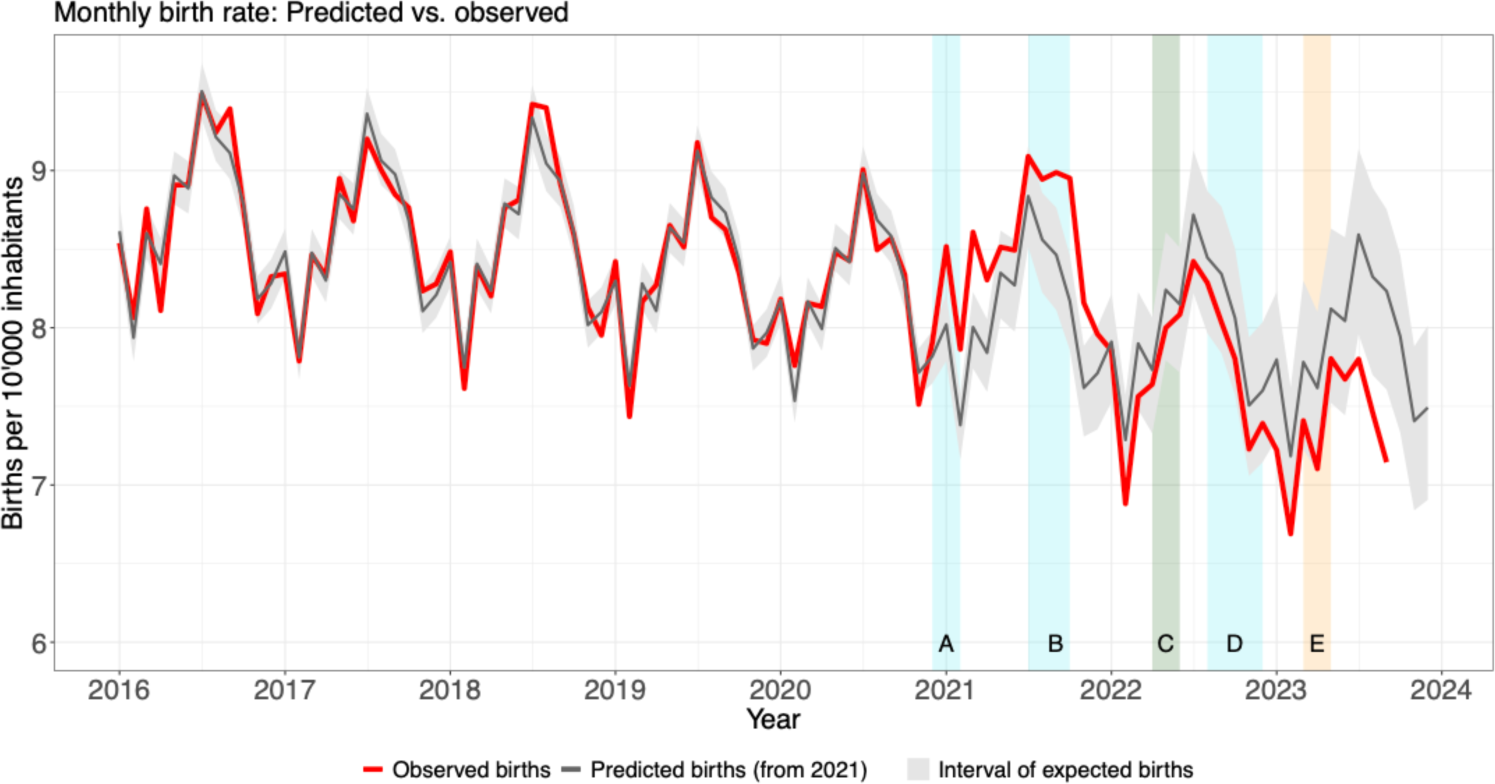
The monthly live birth rate in Switzerland from 2016 to 2023 (red line) and the predicted births (grey line and area) considering 2016 to only 2020 (=pre-pandemic time) as reference births rate. A: The time period 9 months after the first COVID-19 wave in Spring 2020 and the associated partial shutdown; B: The time period 9 months after the second COVID-19 wave in Fall and Winter 2020/2021 and the associated partial shutdown; C: The time period 9 months after the share of double-vaccinated young persons aged 20-40 years surpassed 30% to 50%; D: The time period 9 months after the large Omicron wave in Winter 2021/2022; E: The time period 9 months after the heat wave in Summer 2022.

Based on the fully anonymized individual data, we can also track the trends up to the end of 2022 for certain subgroups of the population (Table 1). The overall pattern (first more births in 2021 and then fewer births in 2022) is also seen in most of the subgroups. The increase in births in 2021 appears to have been slightly stronger in second parities, and slightly weaker in mothers aged <30 years. In Italian-speaking Switzerland and in the case of third and more parities, the increase in 2021 hardly seems to have taken place. The decline in births in 2022 appears to be slightly more pronounced and/or longer in German-speaking Switzerland than in Latin Switzerland, and slightly more pronounced among Swiss mothers, mothers >30 years old, and among non-primiparous women.

## Discussion

We follow population trends in the birth rate in Switzerland almost up to the present and place the latest developments during the COVID-19 pandemic in a historical context. The birth rate in 2022 was the lowest it has been since the 1870s, and it seems the trend is continuing in 2023. The latest decline had already begun 1-2 years before COVID-19. Previous pandemics (1890, 1918, 1920, 1957) had each led to a temporary decline in the birth rate around 9 months after the peak of these outbreaks. With Covid-19, this appears more complex. The immediate shock of the global outbreak has not left a negative mark on births in Switzerland. However, during and shortly after the first two pandemic waves and partial shutdowns in 2020, there were more conceptions and thus excess births in 2021, in all available subgroups except Italian-speaking Switzerland, and somewhat more pronounced among >30-year-old mothers and second parities. Possible reasons for the mini-boom include: The increased time at home during the shutdowns has – planned or not – led to more conceptions which has brought pregnancies forward; the corona virus was still circulating too infrequently in this 1st phase of the pandemic to have a negative impact on pregnancies or fertility; at the end of the waves and shutdowns, the perceived end of the pandemic threat could have led to an optimistic mood and thus also more conceptions. The subsequent decline in births from January 2022 was stronger than the increase in births in 2021. The first part of the decline in 2022 is most likely due to a negative rebound from the advancement of births in 2020/2021 and deliberately postponed pregnancies due to the start of the vaccination program. The second part of the decline in 2022 is associated with conceptions during the big Omicron wave in the winter of 2021/2022, when many people in Switzerland fell ill. In addition, prices have been rising and real wages falling since 2021, the global political situation has become more unstable, and a general change in values regarding the willingness to have children may also be underway.

The short-term shifts in the Swiss birth rate described above are remarkable and show both similarities and differences to other countries. In contrast to other countries (Sobotka et al., 2023), the initial pandemic shock in Switzerland at the beginning of 2020 did not lead to marked drop in the birth rate 9 months later at the end of 2020. The subsequent double-peak increase in 2021, nine months after each of the two pandemic waves and shutdown phases in 2020, also occurred with varying magnitude in other countries. However, the short-term boom was less pronounced or even absent in neighboring countries (Germany, Austria and Italy) (Sobotka et al., 2023). Only France recorded similar increases in 2021 than those reported here. The sharp decline during 2022 is also observed in many other countries, including France, Germany, and Austria (DESTATIS, 2023; Insee, 2023; StatistikAustria, 2023). However, the extent of the short-term ups and downs appears to have been particularly pronounced in Switzerland compared with other countries (Sobotka et al., 2023). The reasons for these country-specific differences still need to be investigated.

Looking back at past pandemics, we can also confirm the temporary decline in births in Switzerland in mid-1919, around nine months after the most severe phase of the “Spanish flu” in Fall and Winter 1918, as in other countries (Bloom-Feshbach et al., 2011; Pomar et al., 2020). This was followed by an increase in births in 1920, although the effects of the end of the war and the end of the pandemic can hardly be separated, even in countries not directly involved in the war (Gaddy & Ingholt, 2023; S.-E. Mamelund, 2012). We also add to the literature by showing that in Switzerland the “Russian flu” in 1890, the strong “Spanish flu” later wave in 1920 and the “Asian flu” in 1957 also led to a short-term drop in births around nine months after the pandemic peaks. In the case of the 1890 Russian flu, the authorities at the time were already aware of the phenomenon (Schmid, 1895), and explained the lack of births around 9 months after the pandemic peak with postponed conceptions as well as infection-related abortions early in pregnancy, which would escape statistics (Schmid, 1895). As far as we know, there are not many comparative studies on birth rates and fertility during these other historical pandemics, but in Switzerland the 1890, 1918-1920, and 1957 pandemics were the strongest before COVID-19 (Staub et al., 2022), and the respective authorities at the time estimated that at least two thirds of the population fell ill during each of these pandemics. This was not the case with the later and weaker pandemics of 1968-70 (“Hong Kong flu”) or 2009 (“Swine flu”), which were not associated with a decline in birth rates in Switzerland.

Usually, crises and pandemics tend to have a negative effect on the birth rate (Lee et al., 2023). The temporary baby boom in 2021 around 9 months after the first two waves of the Covid-19 pandemic and the associated shutdowns in 2020 is therefore rather surprising. There are certainly several reasons for this. On the one hand, thanks to the strong public health interventions (including two partial shutdowns), the number of infections among the population was still relatively low in 2020. This assumption is supported by seroprevalence studies carried out at the time, which showed that after the first wave in spring, around 12% of the population had antibodies in their blood, and this level only increased to around 25% during the second wave in autumn/winter 2020 (West et al., 2020). This means that the risk of infection-related reduced fertility or abortions in Switzerland was probably still rather low in 2020. During the shutdowns, people obviously spent more time at home, which could have improved the work-personal life balance and increased the frequency of intercourse. This could explain the increase in both planned and unplanned (Lewis et al., 2021) pregnancies, perhaps also in the sense of bringing planned pregnancies forward. We also see that the two increases in the number of conceptions in 2021 lasted for a short time after the measures were lifted. The end of the partial shutdowns and the perceived threat of a pandemic may have led to a mood of optimism when it comes to family planning. However, the societal and health impacts of the first two COVID-19 waves and the associated shutdowns in 2020 were certainly not the same for everyone in Switzerland, and this heterogeneity would need to be investigated in more in-depth studies.

The start of the decline in births from January 2022 is associated with conceptions from March 2021. Again, there are various possible reasons, which are not necessarily exclusive. If there was a forward-bringing of pregnancies in 2020 as described above, this could have resulted in a negative rebound from Spring 2021. In addition, the vaccination campaign officially started at the beginning of 2021, but young people without pre-existing conditions were not officially eligible for the vaccination until May 2021 (and the vaccination rate among young people increased only slowly thereafter). This could have led to pregnancies being deliberately postponed until after vaccination, as has been suggested for other countries. At present, the evidence is rapidly growing that the coronavirus vaccination cannot be directly and biologically linked to the decline in births and fertility (Wang et al., 2023; Zaçe et al., 2022). This unanimous opinion is supported by a growing number of studies with different designs on male/female fertility (Aharon et al., 2022; Ba et al., 2023; Barda et al., 2022; Gonzalez et al., 2021; Morris, 2021; Reschini et al., 2022; Yang et al., 2023; Yildiz et al., 2023) and miscarriage (Yland et al., 2023; Zauche et al., 2021), including already some review articles (Rimmer et al., 2023; Wesselink et al., 2022; Zhang et al., 2023; Zhu et al., 2023). Several studies report changes in the duration of menstrual cycle length following COVID-19 vaccination (Alvergne et al., 2023), but these changes are small (+/-1 day) and resume in the next cycle, thus not threatening fertility (Alvergne, 2023). In the case of Switzerland, the timing also speaks against such a direct effect of vaccination on birth rates: The decline in births at the beginning of 2022 and thus in conceptions from spring 2021 was already ongoing for several months before young people in Switzerland were able to be vaccinated, and before >30% of the 20-40 age group were double vaccinated in July 2021.

Where there is a relatively large body of evidence, however, is how single and multiple COVID-19 infections can harm male and female fertility (Aksak et al., 2022; Chen et al., 2021; Harb et al., 2022; Hosseini et al., 2023; Martinez et al., 2023; Patel et al., 2021; Saadedine et al., 2023; Wesselink et al., 2022). And this could be one of the reasons for the decline in births in the second half of 2022. The conception of these pregnancies goes back to the fall/winter of 2021/2022 when the long and massive first Omicron wave hit the Swiss population, and this time a large proportion of the population fell ill. The fact that the birth rate continues to fall in 2023 may also be related to the increasingly uncertain economic situation in Switzerland. Although unemployment never increased massively during the Covid-19 crisis, the consumer price index has been rising steadily since 2021, which is currently leading to a noticeable loss of real wages (Supplementary Figures S7 & S8). The increasingly frequent heatwaves could also have a certain influence, as studies have already linked such extreme climatic events to reduced fertility (Barreca et al., 2018). The increasingly uncertain political situation in the world, for example with the start of the war in Ukraine in February 2022, could also have an impact on family planning.

Finally, life planning decisions are central (Tasneem et al., 2023). Birth rates had already been falling slightly for a few years before the Covid-19 pandemic in Switzerland. In 2018, around 9% of young people said in a nationwide survey that they did not want to have children (Schweiz. Bundesamt für Statistik BFS, 2018). Vasectomies are also on the increase. And it is possible that a change in values is taking place at a societal level, and that some young people simply want to have fewer or no children now, or in general (Guzzo & Hayford, 2023). A possible explanation for the decision to not have children is also concerns about climate change, but also economic or general insecurity could be further reasons (Danny, 2023; Dillarstone et al., 2023)

The present study has several limitations. Firstly, it operates at the population level, which means that there is a lack of in-depth variables in the individual data. For example, at the individual level, nothing can be said about the socio-economic background or other relevant subgroups of the population. Secondly, the natural time lag between conception and birth makes it difficult to interpret the birth rate. The life decisions that are made now are not reflected in the birth rate only about 9 months later. In addition, this temporal lag must be approximated on an ecological level by means of a 9-month distance, as the exact date of birth of the children is not accessible for data protection reasons, which would allow the time of conception to be narrowed down more precisely by means of the gestational age. Thirdly, most official statistics are based on live births only. While stillbirths are fortunately rather rare in Switzerland today (<4.5/1000 yearly between 2020 and 2022), this was not the case a few decades ago, let alone at the end of the 19th century. Unfortunately, there are no monthly stillbirth figures for Switzerland for the time 1871 to 1987 that would allow these cases to be considered in our analyses. Fourth, there is no central database in Switzerland in which new pregnancies are registered. This means that the number of births cannot be verified with reliable statistics on new pregnancies or abortions. Finally, the possible reasons given for the ups and downs in the birth rate are based on temporal associations, and a causal relationship between these is not necessarily the case and has not been investigated. In addition, the reasons are certainly multifactorial and other as yet unknown factors may also play a role. It must be left to more in-depth studies to shed more light on this.

Societies and even specific subgroups such as young people are by no means a homogeneous group. That diversity potentially explains in different ways the ups and downs in births. The present study described the rather surprising and short-term shifts in the birth rate in Switzerland in the last two to three years at the population level. More in-depth studies of different designs and at different levels must now follow to better understand these more or less active fertility decisions.

## Data Availability

The data used in this study are publicly available from the Swiss Federal Statistical Office or can be ordered from them in a fully anonymised form.

## Author statements

### Acknowledgements

We would like to thank the Swiss Federal Statistical Office for providing the fully anonymized data 1987-2022 and further information, as well as Marcel Zwahlen and Svenn-Erik Mamelund for their helpful comments.

### Funding

This work was supported by the Swiss National Science Foundation under Grant 197305.

### Competing interests

The authors declare no competing interests.

### Ethics committee approval

Ethics approval was not required for the reuse of these publicly available and fully anonymized Federal authority data.

### Data availability statement

The aggregated datasets presented in this study and the codes can be found in online repositories:

Github: https://github.com/KaMatthes/birth_trend_Switzerland

Zenodo: https://doi.org/10.5281/zenodo.10277091

## Supplementary Material

**Supplementary Figure S1:**
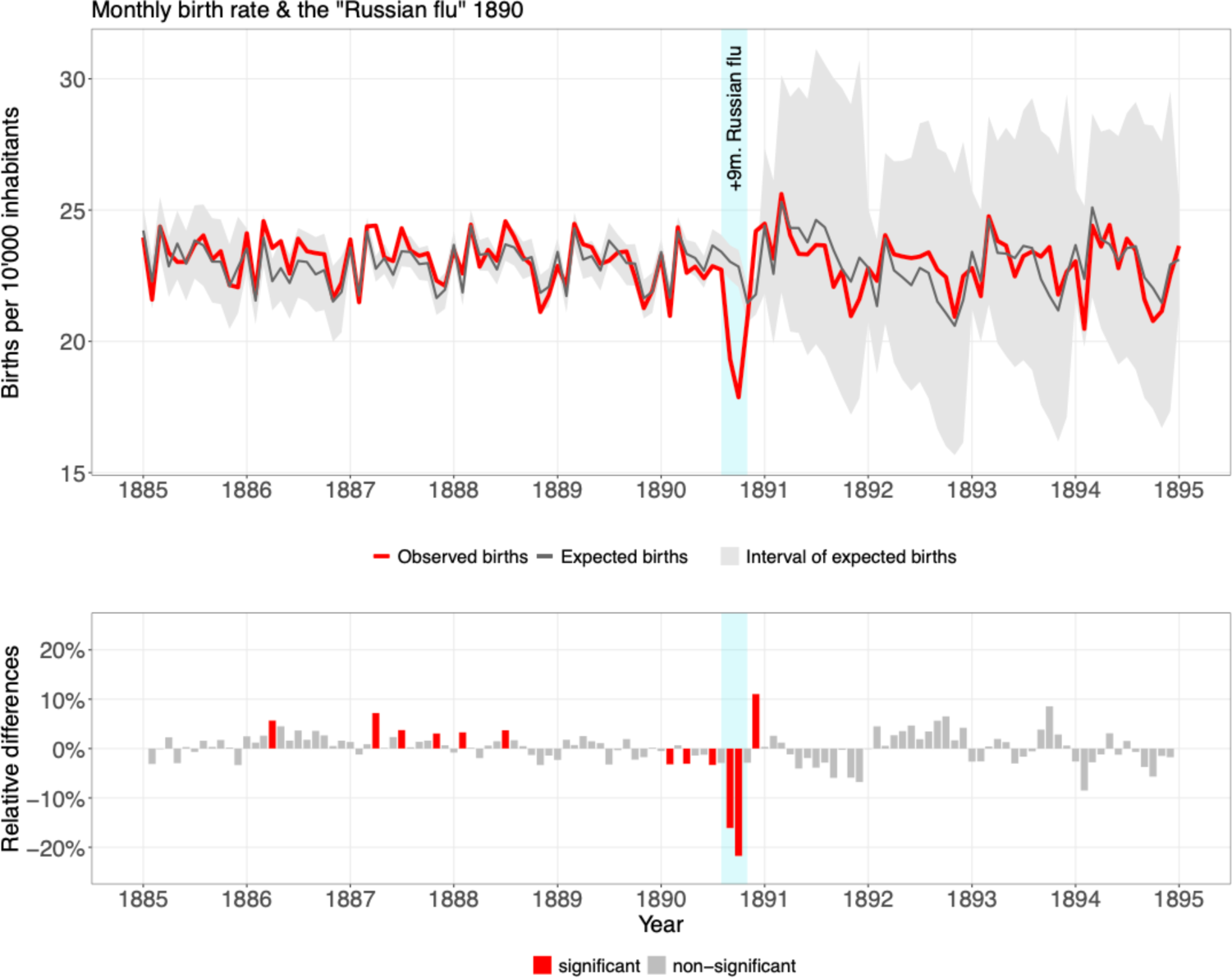
The monthly live birth rate in Switzerland from 1885 to 1895. Top: The observed values (red) compared with the expected values (grey, with interval). Below: The same numbers converted into relative differences, expressed in per cent (red=excess and lower birth rates). Cyan bar: Time period 9 months after the peak of the “Russian flu” in January 1890.

**Supplementary Figure S2:**
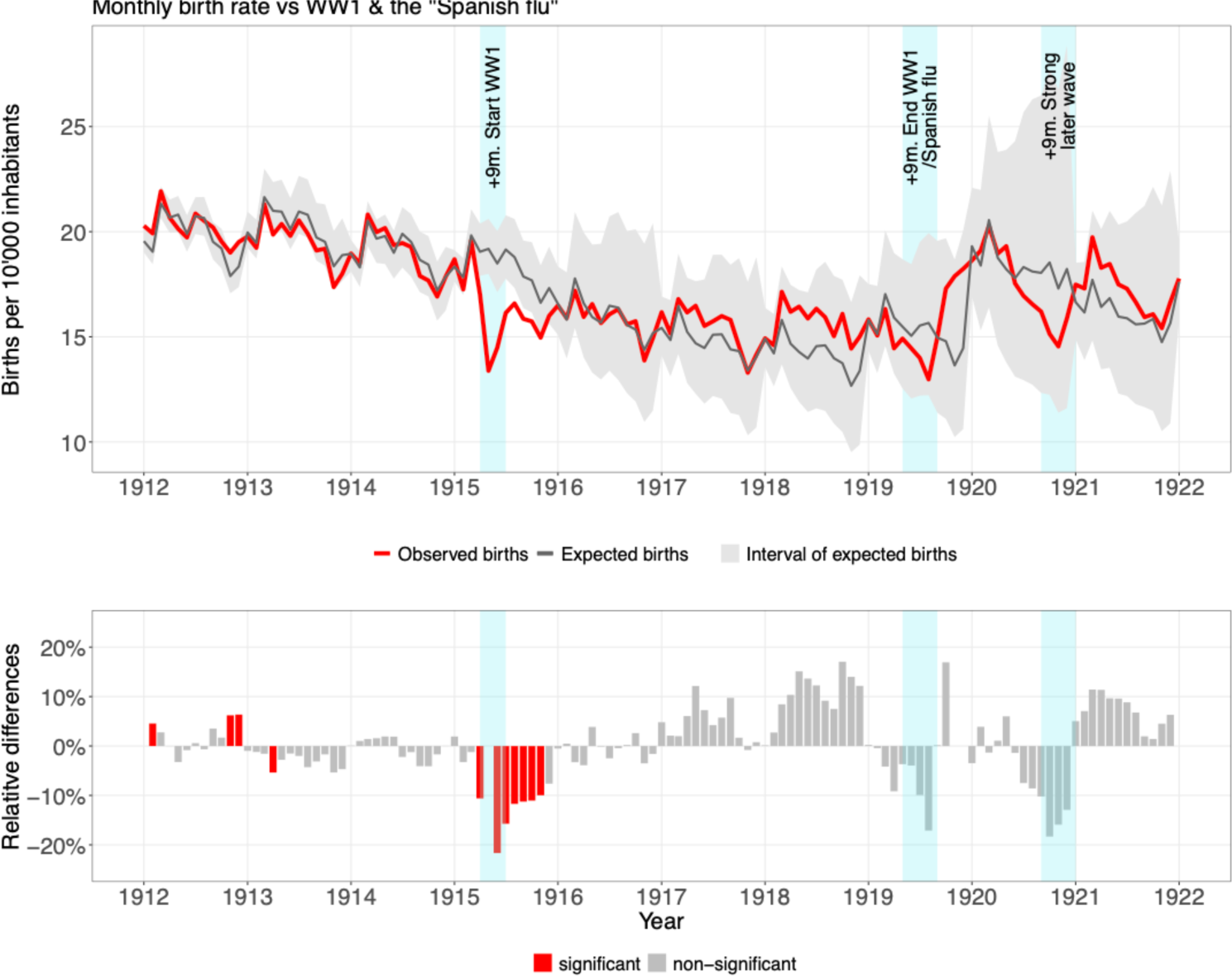
The monthly live birth rate in Switzerland from 1912 to 1922. Top: The observed values (red) compared with the expected values (grey, with interval). Below: The same numbers converted into relative differences, expressed in per cent (red=excess and lower birth rates). Cyan bars: Time period 9 months after the start of World War 1 and mobilization of troops in August 1914; Time period 9 months after the strong Fall/Winter wave of the “Spanish flu” 1918 and the worsening nutritional situation towards the end of the war; Time period 9 months after the strong later pandemic wave in February 1920.

**Supplementary Figure S3:**
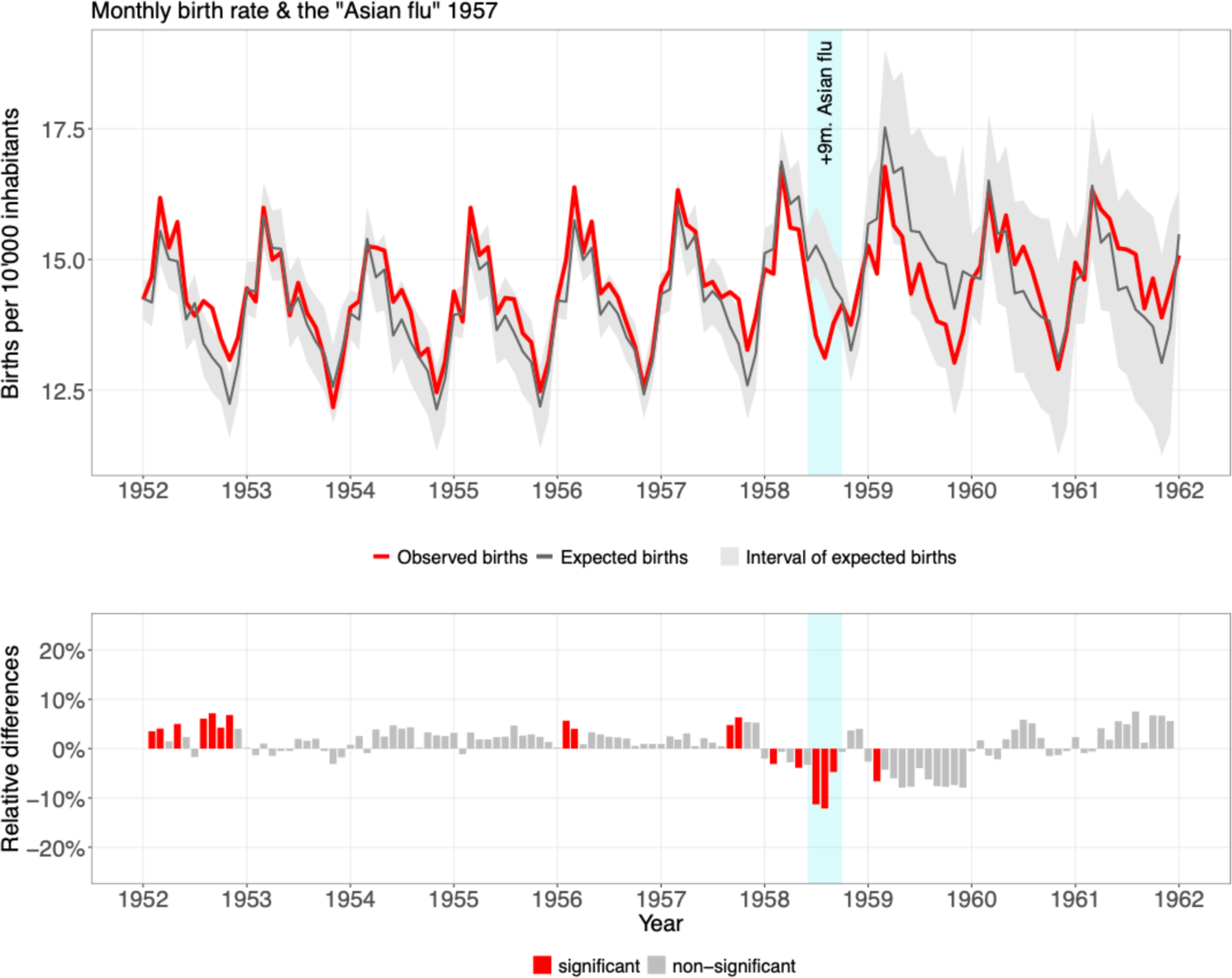
The monthly live birth rate in Switzerland from 1952 to 1962. Top: The observed values (red) compared with the expected values (grey, with interval). Below: The same numbers converted into relative differences, expressed in per cent (red=excess and lower birth rates). Cyan bar: Time period 9 months after the “Asian flu” in Fall 1957.

**Supplementary Figure S4:**
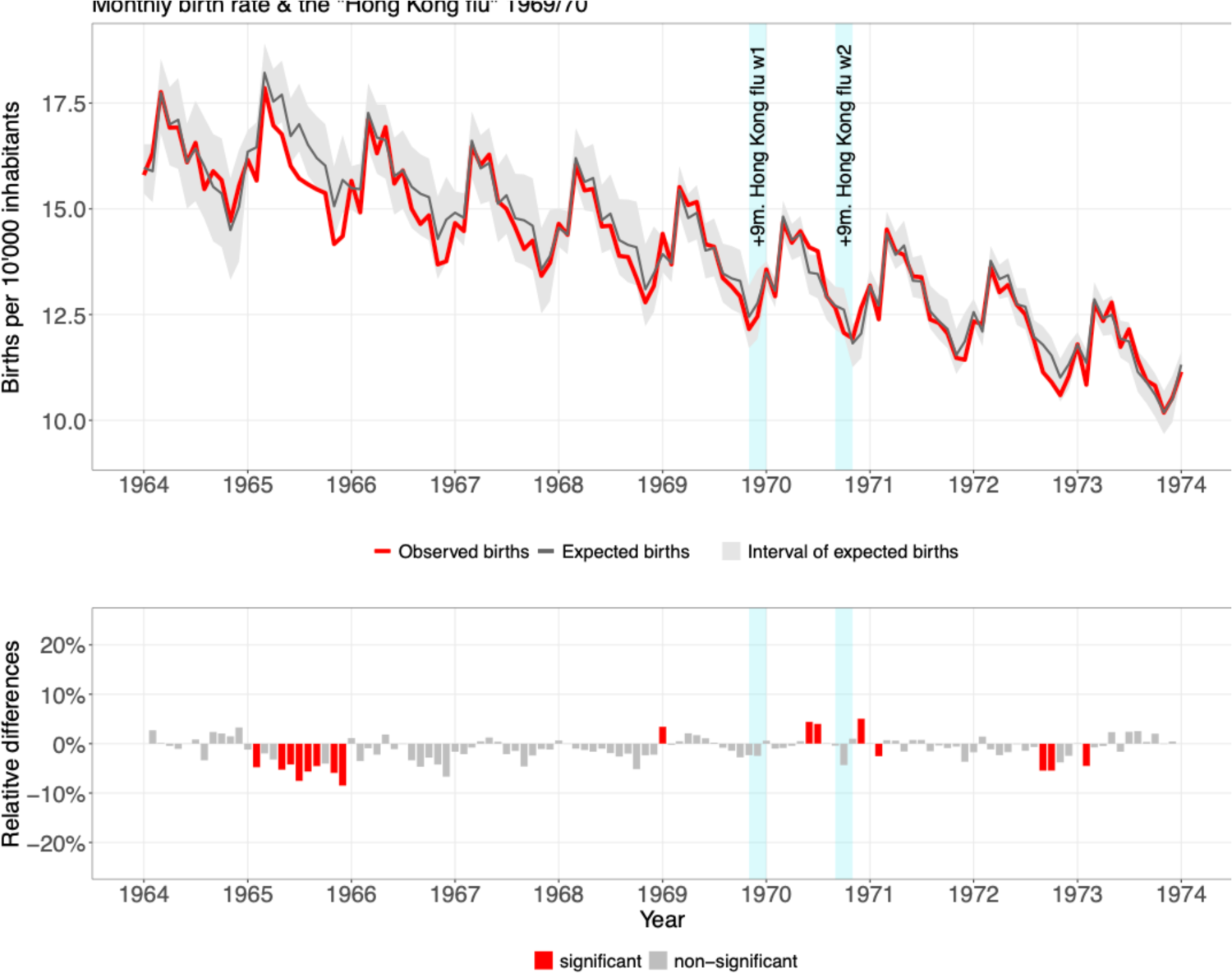
The monthly live birth rate in Switzerland from 1964 to 1974 Top: The observed values (red) compared with the expected values (grey, with interval). Below: The same numbers converted into relative differences, expressed in per cent (red=excess and lower birth rates). Cyan bars: The time periods 9 months after the two waves of the “Hong Kong flu” in Winter 1968/1969 and in Winter 1969/1970.

**Supplementary Figure S5:**
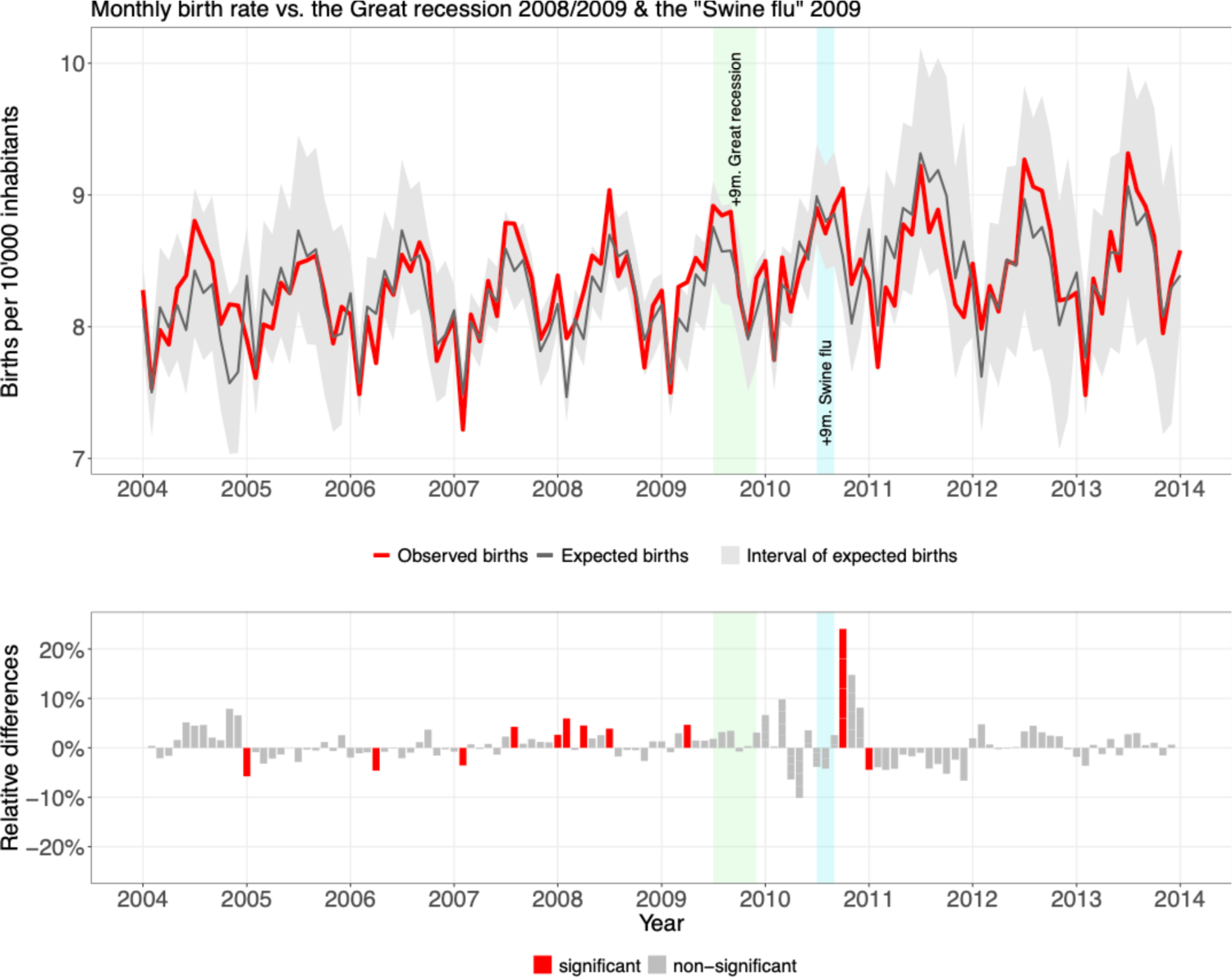
The monthly live birth rate in Switzerland from 2004 to 2014. Top: The observed values (red) compared with the expected values (grey, with interval). Below: The same numbers converted into relative differences, expressed in per cent (red=excess and lower birth rates). Cyan bar: The time period 9 months after the wave of the Swine flu in Fall and Winter 2009. Green bar: The time period 9 months after the Great Recession economic crises at the end of 2008 and the beginning of 2009.

**Supplementary Figure S6:**
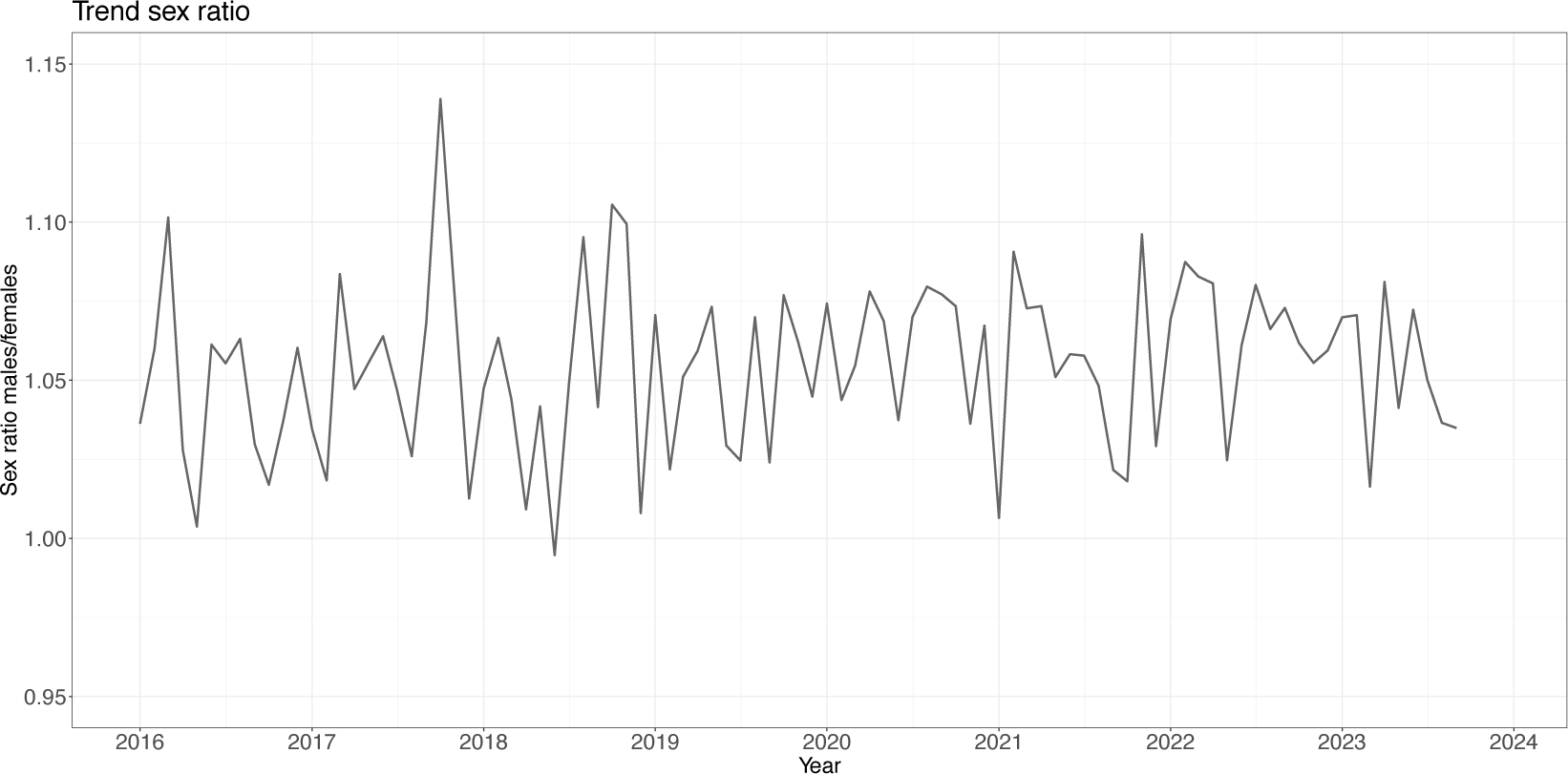
The stable monthly trend in the sex ratio of all live births in Switzerland 2016 to 2023.

**Supplementary Figure S7:**
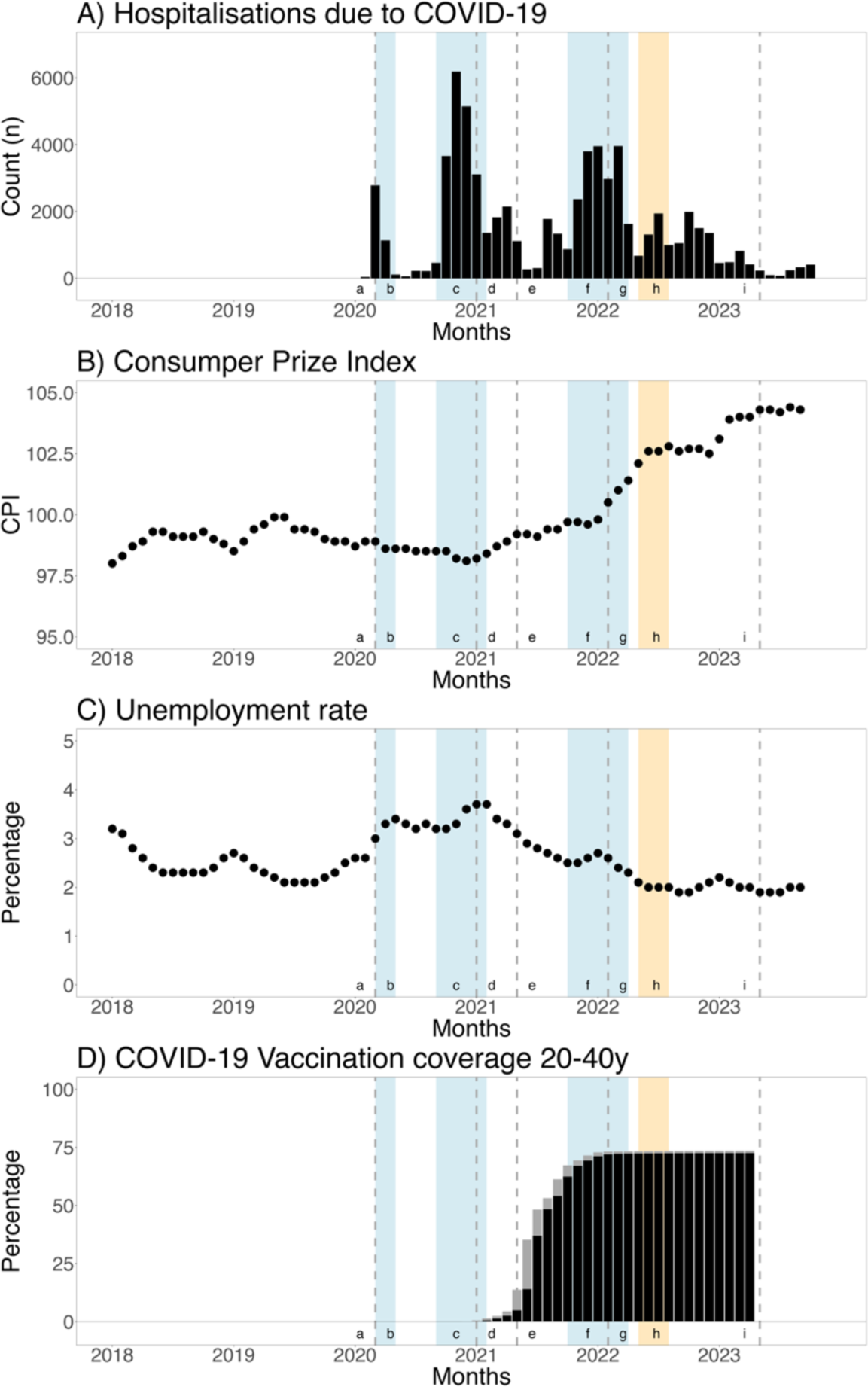
To illustrate the context: A) The different waves of the COVID-19 pandemic in Switzerland, shown here using monthly hospitalisations; B) The monthly trend in the consumer price index in Switzerland; C) The monthly trend in the unemployment rate in Switzerland; D) The vaccination rate among 20–40-year-olds in Switzerland as of 2021 (grey=vaccinated once, black=vaccinated twice). a) The WHO declares the COVID-19 pandemic (dashed line); b) the first wave in spring 2020 (blue bar); c) the second wave in autumn/winter 2020 (blue bar); d) the official start of the vaccination campaign in Switzerland; e) around 10% of 20-40-year-olds have been vaccinated once (dashed line); f) the first wave of the Omicron coronavirus (blue bar); g) the start of the war in Ukraine (dashed line); h) the heatwave in summer 2022 (yellow bar); i) the WHO declares the end of the pandemic (dashed line).

**Supplementary Figure S8:**
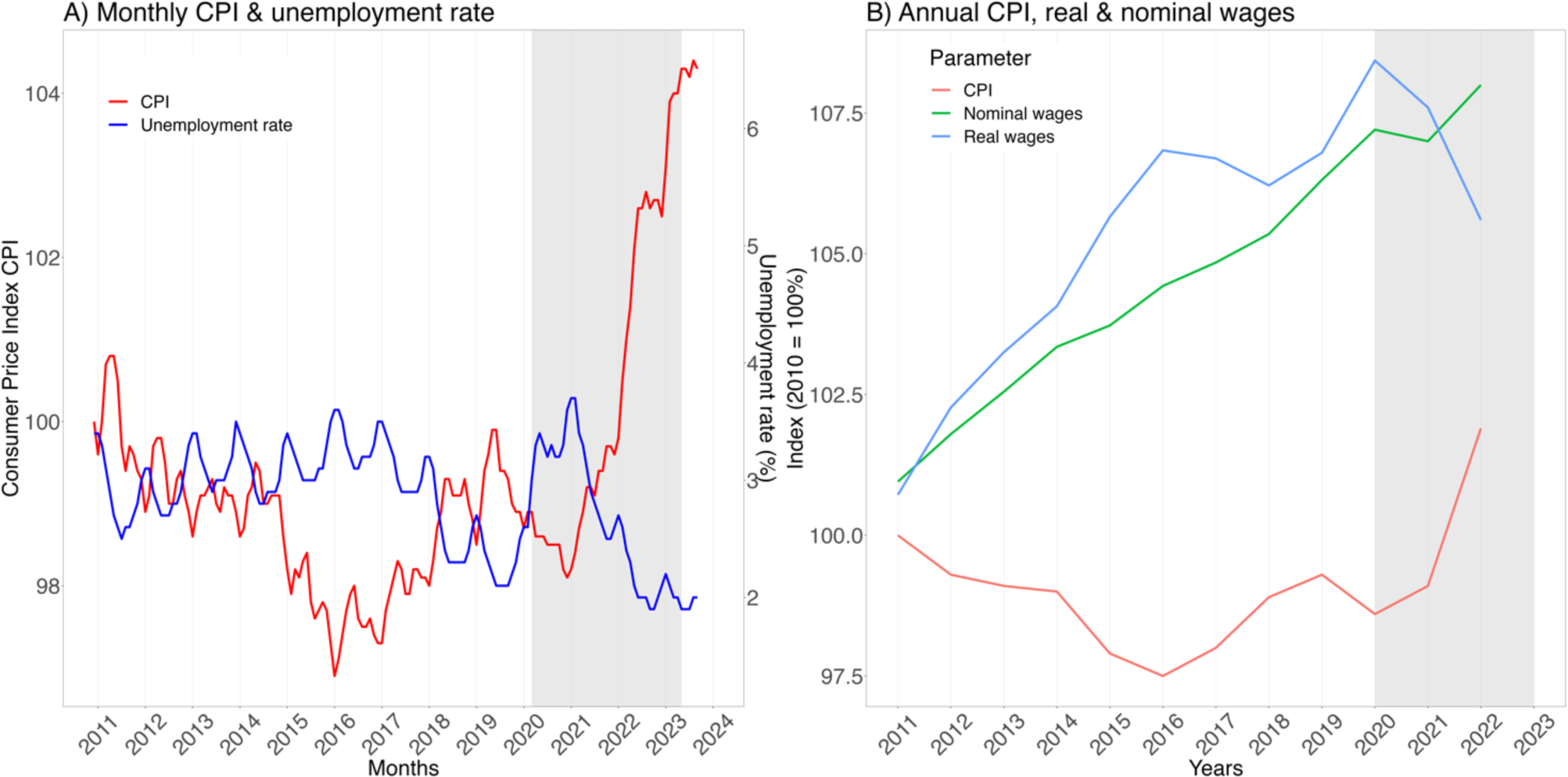
To illustrate the economic context: A) Monthly consumer price index (CPI) and unemployment rate in Switzerland 2011 to 2023; B) Annual CPI, nominal and real wages in Switzerland 2011 to 2022. (Grey bar=time period of the COVID-19 pandemic)

**Supplementary Table S1:**
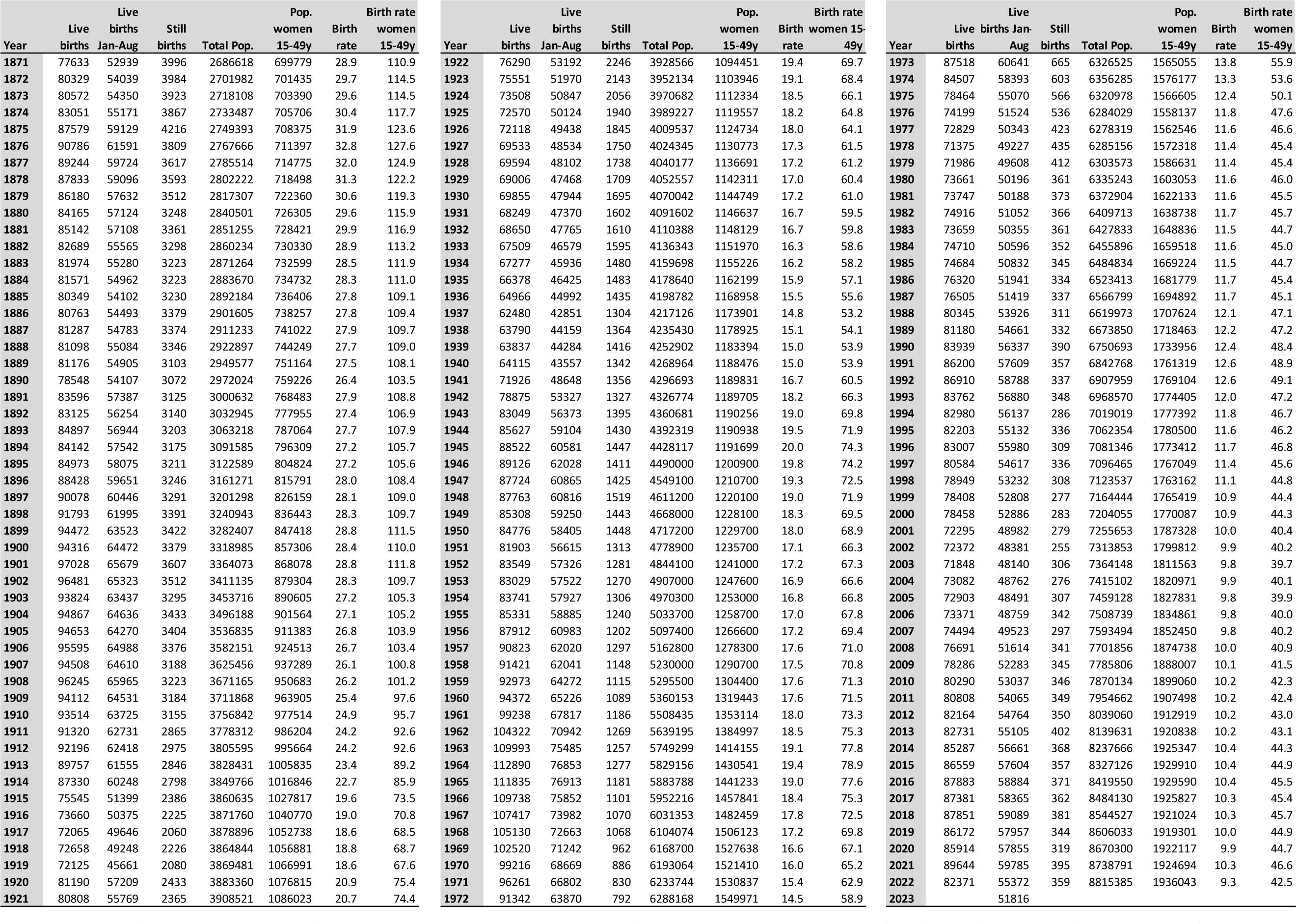
The annual data on births in Switzerland since 1871.

## Notes

### Competing Interest Statement

The authors have declared no competing interest.

### Author Declarations

According to the Human Research Act HRA, no ethical approval is required when working with fully anonymized government data.

### Summary of Updates

The analysis and results are unchanged, the wording has been improved.

